# Each patient is a research biorepository: Informatics-enabled research on surplus clinical specimens via the Living BioBank

**DOI:** 10.1101/2020.09.25.20199679

**Authors:** Alexander V. Alekseyenko, Bashir Hamidi, Trevor D. Faith, Keith A. Crandall, Jennifer G. Powers, Christopher L. Metts, James E. Madory, Steven L. Carroll, Jihad S. Obeid, Leslie A. Lenert

## Abstract

The ability to analyze human specimens is the pillar of modern-day translational research. To enhance the research availability of relevant clinical specimens, we developed the Living BioBank (LBB) solution, which allows for just-in-time capture and delivery of phenotyped surplus laboratory medicine specimens. The LBB is a system-of-systems integrating research feasibility databases in i2b2, a real time clinical data warehouse, and an informatics system for institutional research services management (SPARC). LBB delivers de-identified clinical data and laboratory specimens. We further present an extension to our solution, the Living µBiome Bank, that allows the user to request and receive phenotyped specimen microbiome data. We discuss the details of the implementation of the LBB system and the necessary regulatory oversight for this solution. The conducted institutional focus group of translational investigators indicates an overall positive sentiment towards potential scientific results generated with the use of LBB. Reference implementation of LBB is available at https://LivingBioBank.musc.edu.

## Statement of the problem

The need for molecular and other data from precisely phenotyped human specimens paramount in translational research. Although a part of the general solution, expansive biobanking of such specimens is not always viable due to the high burden and expense of biobank establishment and maintenance [1]. Although not a complete replacement, just-in-time capture of specimens from standard-of-care surplus sources can support many use cases of traditional biobanking, if this can be accomplished with minimal disruptions to the clinical laboratory’s mission. Examples of existing platforms that provide just-in-time specimen capture functionality include: (i) Harvard’s Crimson system, which combines its Informatics for Integrating Biology and the Bedside (i2b2) [2] with a link to its laboratory specimen processing system [3, 4]; and (ii) the Vanderbilt BioVU system, which operates more like a traditional biobank with storage of a broad range of specimens [5, 6].

Living BioBank (LBB) seeks to extend the model developed in Crimson from its proprietary platform to a more generally applicable solution and to extend the approach across research networks. In addition, a limitation of the Crimson system is the necessity for the specimens and patient data to be identified to the end user. The specimen recruitment process involves identified chart review by investigators to accept or reject the available specimen. This typically requires a higher degree of regulatory oversight than if the specimens were to be de-identified and clinical abstraction limited to just a predefined phenotype. As a result, documentation and time requirements for such studies increase. A more streamlined solution would allow for a precise electronic phenotype to guide the surplus specimen capture without the need to validate every individual patient’s record.

In this case report, we describe the Medical University of South Carolina Biomedical Informatics Center LBB solution for phenotyped surplus specimen capture that integrates a number of institutionally adopted informatics systems to deliver some functionality of just-in-time biobanking in a reduced regulatory burden environment. We demonstrate the use of LBB in an IRB-approved Living µBiome Bank study protocol that further seeks to augment the phenotype with phenotyped specimen microbiome data and limited de-identified clinical data to support microbiome data interpretation. We assess the feasibility of the LBB solution in terms of the accuracy of e-phenotypes and the time required for delivery of a set of specimens. We also report on evaluation of user acceptance of the solution.

## Technical description of the Living BioBank

**Figure 1A** outlines the LBB solution players, systems, processes, and deliverables. Although many electronic phenotyping systems may suit the purpose, the current implementation of the LBB (https://LivingBioBank.musc.edu/) uses i2b2 as an entry point for specimen requests. A request is specified by an investigator seeking surplus laboratory materials. An honest data broker retrieves and reviews the i2b2 phenotype with the investigator to ensure the best match of the specified inclusion/exclusion characteristics to the intended phenotype. The query is then used to match patients who have laboratory testing ordered and to deliver just-in-time surplus specimen availability reports to laboratory honest brokers. The primary specimen source for the LBB is the MUSC Pathology and Laboratory Medicine phlebotomy operations. These specimens are available for capture after clinical testing is completed and just prior to when the specimen is normally discarded. At this point all identifiers are stripped from the physical specimen, and the specimen is no longer considered human subject derived material. The laboratory honest brokers fulfill the requests by delivering a collection of phenotyped specimens to investigators. Thus, the final LBB deliverables include the reviewed i2b2 phenotype (query) and phenotyped specimens. Specimens are only collected when matching to a specified phenotype request and become surplus and no longer needed for provision of care.

**Figure 1.**
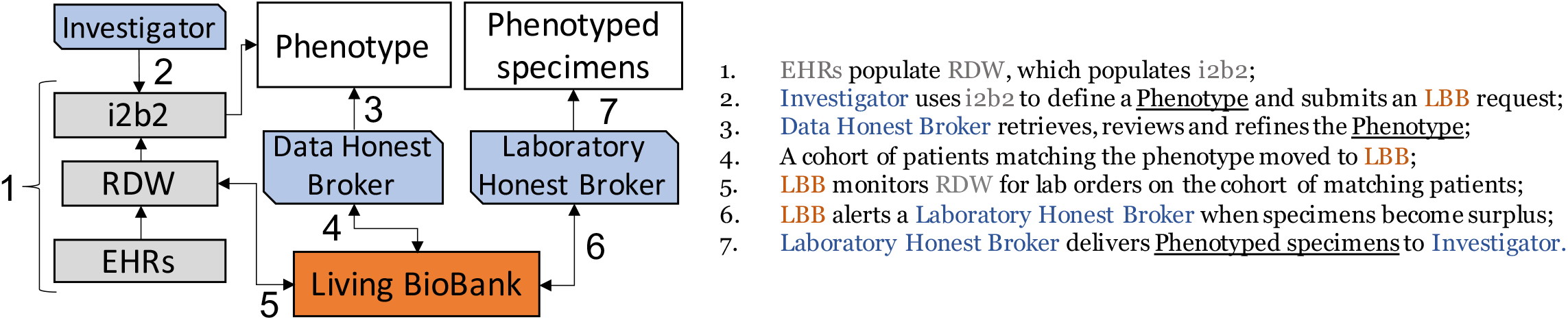
The Living BioBank architecture integrates phenotyping, data warehouse, and honest data and laboratory broker services.

A walk-through of the LBB user and honest broker interfaces is available as **Supplementary data**.

### Regulatory status of the LBB

The development of LBB proceeded under regulatory consultation with the MUSC Institutional Review Board (IRB). LBB was determined to be a process and thus exempt from review. From the MUSC IRB standpoint the phenotyped specimens do not constitute human subjects derived materials and therefore are exempt from regulations regarding informed consent. Further the phenotypes used for specimen capture are deemed generic enough to not constitute identifiable data. LBB users are required to abide by a Data Use Agreement, which explicitly prohibits re-identification of the subjects. In conjunction with the LBB process to capture phenotyped specimens, the users may use institutional data request services to obtain linked clinical variables. Such requests require proper oversight, including IRB-approved protocols, which are reviewed by data honest brokers to determine compliance and allowable clinical data elements to be delivered. For our study, the MUSC IRB has determined that the surplus specimens delivered via LBB proces comprise de-identified non-human subjects material. However, each study would still have its own IRB review, and if the requested data or samples were deemed potentially identifiable, HIPAA would have to be addressed either via a limited data set agreement or a HIPAA waiver. This may happen, for example, if the phenotyped specimen capture occurs in such a short time-span that, even if the specimen date is not explicitly provided, it may be viewed as implicitly available, thus potentially being interpreted as a limited identifier in the data.

## Living µBiome Bank for phenotyped microbiome data requests

As an illustration of a practical use case of the LBB solution, we present an NIH NCATS-funded research study protocol, the Living µBiome Bank (LµBB), developed to deliver phenotyped specimen microbiome data using LBB solution (**Figure 2A**). The LµBB protocol was approved by IRB (Pro00079660) to deliver to the users processed phenotyped specimen microbiome data and an additional limited set of non-PHI clinical variables from the medical records to facilitate data interpretation. The institutional research services necessary for LµBB request fulfillment are managed by integration with the Service, Pricing, and Applications for Research Centers, or SPARCRequest (SPARC) system, which is an interoperable, searchable research-resource electronic storefront that supports research services operations [7]. LµBB uses the LBB solution to remove the user from physical access to the specimen by capturing the phenotyped specimens and processing them to derive deliverable microbiome assay data. The LµBB finds suitable specimens for many possible microbiome studies at MUSC Infection Control active surveillance programs. At MUSC, surveillance for methicillin-resistant *Staphylococcus aureus* (MRSA) and vancomycin-resistant *Enterococci* (VRE) generate over 15,000 tests for each pathogen annually. Recently, COVID-19 testing nasopharyngeal swabs have become available as a specimen source. The associated clinical data is available to aid in interpretation of the microbiome data and is delivered de-identified by the data honest broker.

**Figure 2.**
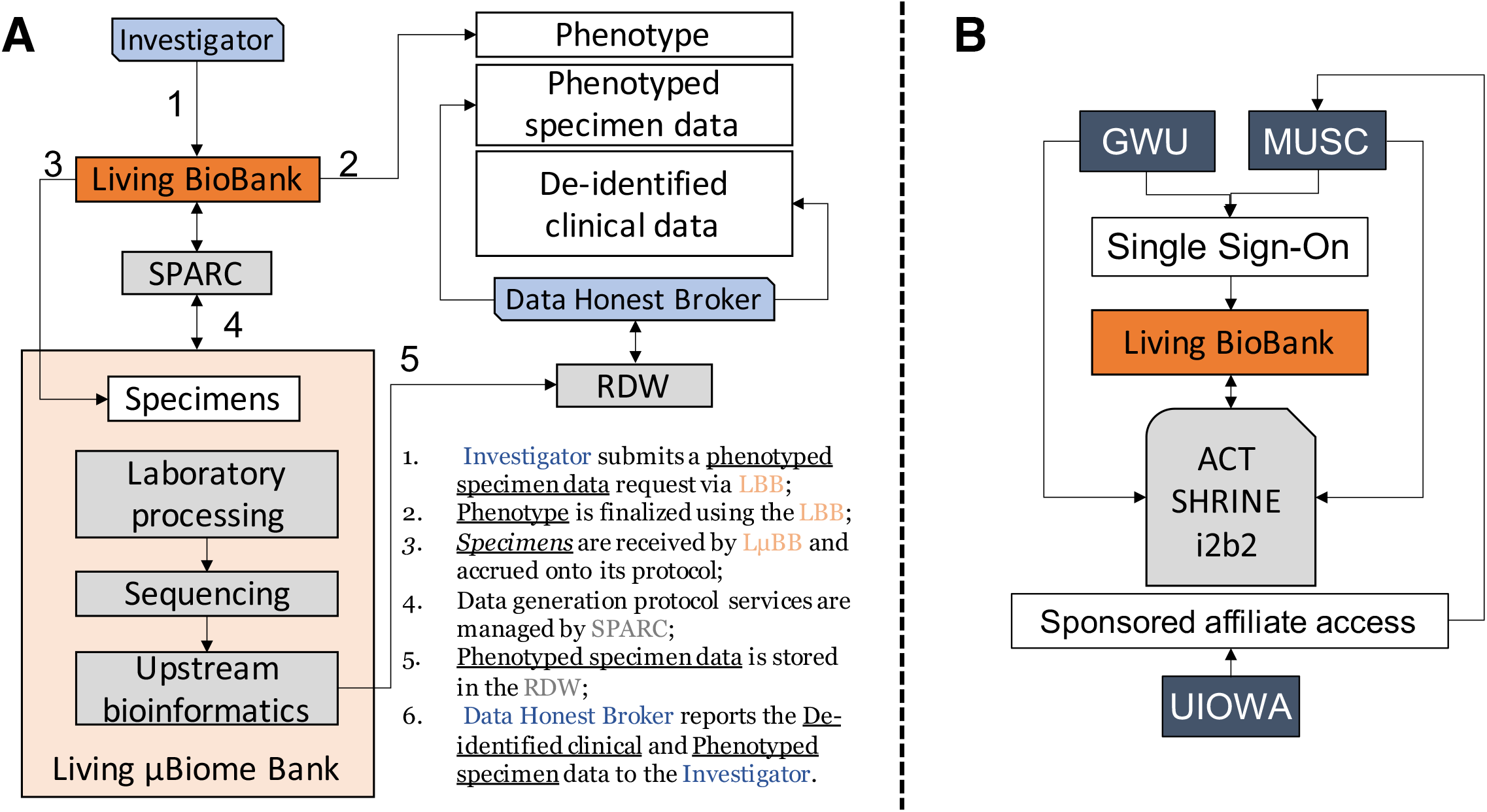
Implementation of the Living µBiome Bank study protocol using LBB solution (A) by integrating institutional research services request system into specimen processing pipeline; and (B) multi-institutional access solution.

### Multi-institutional access to the LµBB

We implemented a model for inter-institutional requests within the LµBB. The working solution presented in **Figure 2B** uses CTSA Accrual to Clinical Trials (ACT) Network Shared Health Resource Information Network (SHRINE) [8] to share the phenotype specification from the requesting institution (e.g., GWU) to MUSC LµBB. Authentication to the LBB application is enabled by InCommon, a widely accepted authentication infrastructure across academic institutions in the USA. Non-ACT SHRINE member sites (e.g., UIOWA) can be credentialed at the LBB host site using institutional affiliate sponsored access processes. Under the network model only the data, not the specimens cross the institutional boundaries, which only requires a Data Use Agreement to operate.

### Additional regulatory considerations for LµBB

LµBB study is granted a waiver of consent by the IRB because it is impracticable to obtain consent given that there is no way to know who would have leftover samples, and most patients would already be gone by the time the samples were pulled for research. To further mitigate ethical research concerns LµBB considers when present the patient’s response to Research Permissions Questionnaire (RPQ) implemented by MUSC. RPQ allows patients to indicate if they ‘agree’ or ‘do not agree’ to research on their surplus materials that would otherwise be discarded [9-12]. By default, opt-out preferences are honored; however, since RPQ responses are available in RDW, the i2b2 the users may request that recruitment only happens for subjects indicating opt-in preferences. Our data indicate among the patients who have been presented with the RPQ 70-75% tend to respond in the affirmative [11, 13]. Because of the adoption of the Common Rule, the function of the RPQ is not required for LBB implementation but is an optional additional feature.

## Validation, user expectations and experiences

We identified 21 individual investigators across MUSC, including College of Medicine, College of Dental Medicine, and College of Nursing, to serve as clinical area experts to conduct a validation focus group study, deemed by the IRB to be quality improvement and exempt. Each expert was offered an opportunity to specify their own phenotype of interest using MUSC i2b2 and asked to abstract 20 retrospective charts from each phenotype to determine if those were a ‘Match’, ‘Mismatch’, or if they were ‘Unsure’ for any reason. Nineteen investigators completed chart abstractions and an exit questionnaire.

### Many investigator defined phenotypes result in reasonable patient matches upon review

The chart match rate to the phenotype varied dramatically across experts (**Figure 3**). Within each clinical area certain e-phenotypes captured the desired cohort better than others. On average, infectious, rheumatic, neonatal, and cancer e-phenotypes performed better whereas psychiatric, gastrointestinal, skin, and pulmonary performed worse at capturing the intended cohort of the clinical experts. We speculate that the domains that did well are inpatient focused clinical areas, which naturally collect more data in the EHR, while the domains that did not perform as well are more outpatient focused. Further study of the factors that contribute to successful e-phenotype specification is thus warranted and will be forthcoming from our group.

**Figure 3.**
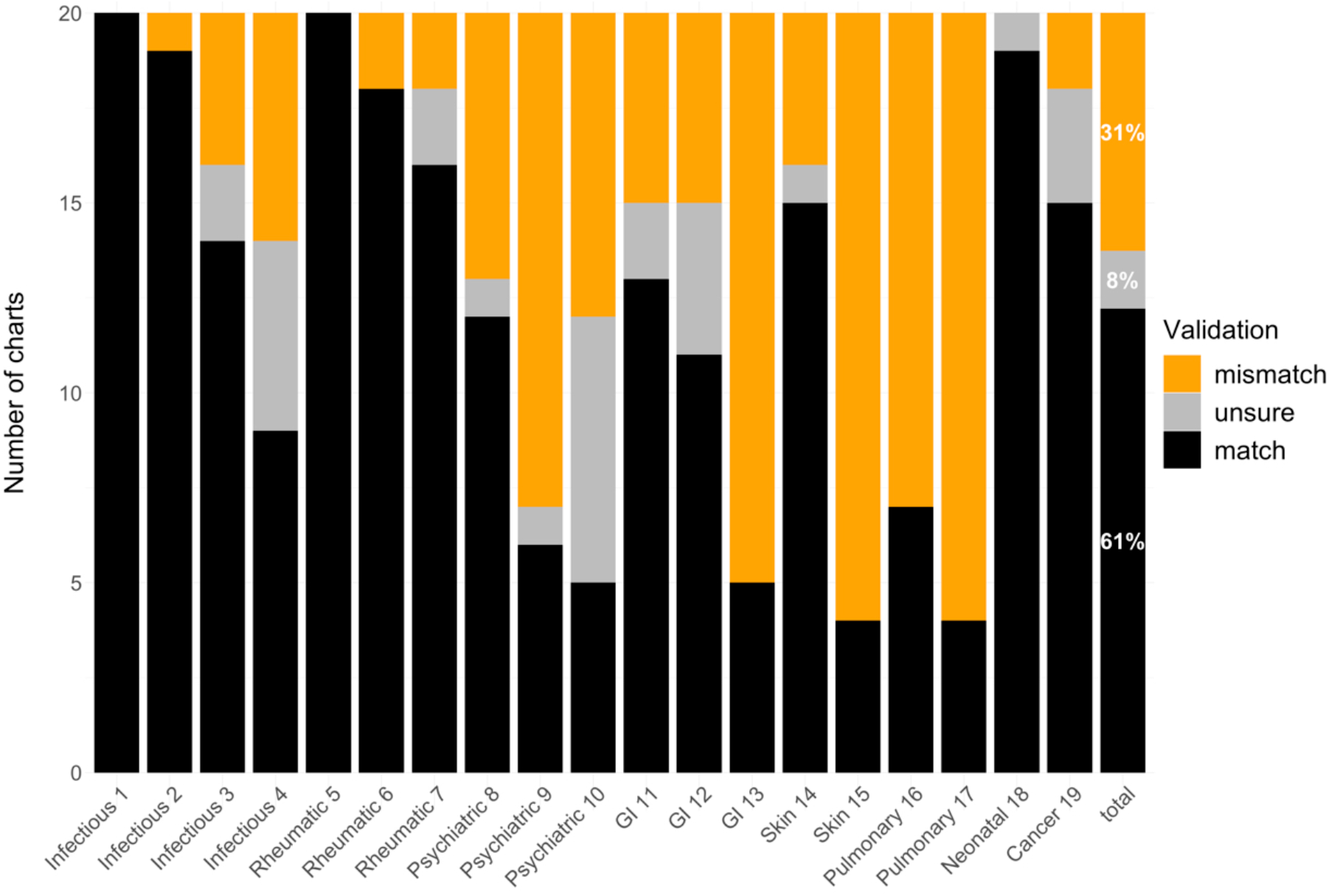
Expert investigator-specified phenotype validation rates suggest feasibility of just-in-time recruitment from i2b2 phenotypes without full chart review.

### Expected specimen accrual times are reasonable and practical for pilot data collection

Using historical data, we have estimated average accrual rates for each phenotype. Most phenotypes yielded accrual rates reasonable for small-scale pilot studies. For instance, as shown in **Table 1**, the majority of specified phenotypes would be able to accrue a sample of twenty specimens in under four weeks. Only a quarter of the specified phenotypes would need more than six months to accrue this sample size; these represent the cases where multi-institutional collaborations are needed. Specific clinical areas where collaborations are needed include pediatric and neonatal phenotypes, rheumatic diseases, and opioid use disorders. Two e-phenotype queries (GI 11 and skin 15) specified pediatric populations, which are excluded from LµBB requests due to limitations of our current implementation of research preferences capture.

**Table 1.** Number of e-phenotypes by expected waiting time (weeks) for accrual of 20 specimens.

| Specimen | <4<br>weeks | 4–26<br>weeks | 26–52<br>weeks | >52<br>weeks |
| --- | --- | --- | --- | --- |
| Blood | 14 | 5 | 0 | 2 |
| Nasal | 6 | 8 | 2 | 3 |
| Gut | 5 | 8 | 2 | 4 |

### Clinical experts expect to trust scientific findings from LBB and are likely to recommend

A REDCap [14] questionnaire has been offered to our investigator focus group to reflect on their experience using i2b2 system and their recommendations for the specimens obtained from LBB (**Table 2**). Responses to the questionnaire were assessed for association with chart-to-phenotype match rates in R (version 3.6.1) [15] using logistic regression models with ordered Likert-scale factors as trends. With the study coordinator and honest broker support provided, most describe i2b2 as easy to use and would be willing to use it again to define a phenotype. Clinical experts who would be willing to use the system again had, on average, a higher match rate, with a positive linear trend (coefficient=0.95, p-value=0.006). Experts generally tended to have confidence in the scientific data from LBB, while six of the experts had some reservations. We also identified a positive linear trend between match rate and the level of confidence in scientific findings from LBB (coefficient=0.55, p-value=0.016). Experts generally reported a willingness to recommend the system to others, with little to no reservation (n=17). The willingness to recommend came with a positive linear relationship with chart match rate (coefficient=0.73, p-value=0.005). More than half of the experts were willing to pay to obtain specimens from the system. Phenotypes specified by this group of experts had an average match rate of 77% (CI 60.7-93.7).

**Table 2.** Questionnaire captures expert reflections on just-in-time biobanking with LBB

| Question and Response | N | Validation match rate (%) |  | Trend test |  |
| --- | --- | --- | --- | --- | --- |
|  |  | Mean (95% CI) | Contrast* | Coefficient | P-value |
| In the future, would you use the i2b2 system again to define a cohort? |  |  |  |  |  |
| Definitely will not use again | 0 |  |  |  |  |
| Probably will not use again | 1 | 35 |  |  |  |
| Probably will use again | 9 | 57.8 (42.1-73.5) | linear | 0.95 | 0.006 |
| Definitely will use again | 9 | 67.2 (45.1-89.4) | quadratic | -0.22 | 0.362 |
| How difficult was the use of i2b2 to define a cohort? |  |  |  |  |  |
| Not at all difficult | 10 | 64.5 (44.4-84.6) |  |  |  |
| Somewhat difficult | 5 | 71 (52-90) |  |  |  |
| Moderately difficult | 3 | 30 (24.3-35.7) | linear | -0.22 | 0.528 |
| Very difficult | 1 | 70 | quadratic | 0.7 | 0.025 |
| Would you trust scientific results that derive from a system like this? |  |  |  |  |  |
| Definitely will not trust | 0 |  |  |  |  |
| Probably will not trust | 6 | 44.2 (30-58.3) |  |  |  |
| Probably will trust | 10 | 70.5 (19.4-107) | linear | 0.55 | 0.016 |
| Definitely will trust | 3 | 63.3 (30-58.3) | quadratic | -0.59 | 0.001 |
| Would you recommend this system to your colleagues? |  |  |  |  |  |
| Definitely will not recommend | 0 |  |  |  |  |
| Probably will not recommend | 2 | 52.5 (18.2-86.8) |  |  |  |
| Probably will recommend | 10 | 52.5 (35.4-69.6) | linear | 0.73 | 0.005 |
| Definitely will recommend | 7 | 75.7 (54.2-97.3) | quadratic | 0.42 | 0.027 |
| Would you pay to obtain specimens from such system? |  |  |  |  |  |
| Definitely will not pay | 1 | 70 |  |  |  |
| Probably will not pay | 8 | 46.2 (29-63.5) |  |  |  |
| Probably will pay | 9 | 77.2 (60.7-93.7) | linear | -1 | 0.037 |
| Definitely will pay | 1 | 25 | quadratic | -0.66 | 0.078 |

\*Only linear and quadratic trends are presented.

## Conclusions and Future Directions

LBB provides a solution for phenotyped surplus specimen capture that is open source (Ruby source code available at https://github.com/living-biobank/living-biobank) and eliminates the need for identified chart review to capture each individual specimen, improving on the Harvard Crimson model. Further the LBB implementation of just-in-time capture of user-requested phenotypes does not require comprehensive biobanking of large volumes specimens without a specific user needing them as in the BioVu model, potentially cutting the costs of human-derived research material provision. Our solution integrates processes across two widely adopted infrastructure systems within the CTSA community, namely SPARC and i2b2. LBB is not a replacement for a proper biorepository, which allows for full access to clinical data for consenting subjects, but can be a useful auxiliary service to accelerate research, as our LµBB study demonstrates. The LBB solution access to phenotypes and phenotyped specimens simplifies regulatory considerations for the users. LµBB further extends LBB to allow for surplus clinical specimens to be processed using high-throughput molecular laboratory techniques (e.g., sequencing) to deliver de-identified phenotyped microbiome and clinical data. This allows the user to deal only with the data rather than the physical specimens. The focus groups study conducted indicated high phenotype match rates and acceptance of scientific potential of the data generated by the LBB process. Further work is needed to scale the system to allow for integrated inter-institutional specimen requests and to understand the factors and best practices in phenotyping to ensure highest possible match rates.

## Data Availability

The authors confirm that the data supporting the findings of this study are available within the article [and/or] its supplementary materials.

## Acknowledgements

The Biomedical Informatics Center personnel tirelessly worked on the LBB: John Clark, Andrew Cates, Katie Kirchoff, Ito Eta, Kayla Glick, Wenjun He, Tami Crawford. AVA and JGP were delegates to 2017 Microbiome Data Science Innovation Labs sponsored by BD2K Training Coordinating Center. We thank Kimberly Snow for providing valuable feedback on clarity and wordsmanship.

## Competing Interests Statement

AVA is a scientific advisory board member for Second Genome, Inc., which has not contributed to this research.

## Funding Statement

AVA, BH, LAL, JSO, KC, and JGP are supported by NIH/NCATS R21 TR002513. AVA and BH are supported by NIH/NLM R01 LM012517. SLC is supported by NIH/NCI 5P30CA138313, 1UM1CA239752 and 1U54CA210962 and by NIH/NIA 1RO1AG055132. The project described was supported by the NIH/NCATS UL1 TR001450.

## Contributorship Statement

LAL and AVA conceived the essential concepts of the manuscript and directed the research, BH and TDF contributed creatively to the implementation of the concept, KAC and JGP contributed to evaluation and network solution, CLM, JEM, and SLC contributed to feasibility of the project from the laboratory medicine perspective. JSO oversaw implementation of essential informatics infrastructure for the project. AVA drafted the manuscript. All co-authors have revised and approved the manuscript.

## Notes

### Author Declarations

The development of LBB proceeded under regulatory consultation with the MUSC Institutional Review Board (IRB). LBB was determined to be a process and thus exempt from review. From the MUSC IRB standpoint the phenotyped specimens do not constitute human subjects derived materials and therefore are exempt from regulations regarding informed consent. Further the phenotypes used for specimen capture are deemed generic enough to not constitute identifiable data.

